# Association between willingness to receive the COVID-19 vaccine and sources of health information among Japanese workers: a cohort study

**DOI:** 10.1101/2021.07.15.21260609

**Authors:** Ko Hiraoka, Tomohisa Nagata, Takahiro Mori, Hajime Ando, Ayako Hino, Seiichiro Tateishi, Mayumi Tsuji, Shinya Matsuda, Yoshihisa Fujino, for the CORoNaWork project

## Abstract

**Background:** It is important to achieve herd immunity by vaccinating as many people as possible to end the COVID-19 pandemic. We investigated the relationship between willingness to receive vaccination and sources of health information among those who did not want to be vaccinated against COVID-19.

**Methods:** This prospective cohort study collected data using a self-administered questionnaire survey. The baseline survey was conducted during December 22–25, 2020, and the follow-up survey during February 18–19, 2021. Participants were aged 20–65 years and worked at the time of the baseline survey (N = 33,087). After excluding 6,051 invalid responses, we included responses from 27,036 participants at baseline. In total, 19,941 people responded to the follow-up survey (74% follow-up rate). We excluded 7,415 participants who answered “yes” to the question “If a COVID-19 vaccine becomes available, would you like to get it?” in the baseline survey. We finally analyzed 12,526 participants.

**Results:** The odds ratio for change in willingness to be vaccinated from “no” to “yes” differed by source of health information. Compared with workers that used TV as a source of information, significantly fewer people who reported getting information from the Internet and friends/colleagues were willing to get the vaccine.

**Conclusions:** It is important to approach workers who do not watch TV when implementing workplace vaccination programs. It is likely that willingness to be vaccinated can be increased through an active company policy whereby the top management recommend vaccination, coupled with an individual approach by occupational health professionals.

**Trial registration:** Not applicable.

## Introduction

In Japan. vaccination against COVID-19 started for about 40,000 healthcare workers on February 17, 2021, in April 2021 for people aged ≥65 years, and on June 21, 2021, for workers in workplaces. Vaccination is voluntary and the decision whether to vaccinate is made by individuals [1]. Obtaining herd immunity through vaccination is important to end an infectious disease pandemic [2]. However, many people are hesitant to get vaccinated, and some refuse vaccination [3]. This vaccine hesitancy has become an important public health issue. In Japan, it is well known that there is a lack of public trust in vaccines [4].

Previous studies revealed factors related to willingness to receive the COVID-19 vaccine. Age, gender, race, and education level were associated with vaccine hesitancy [5-7]. People are required to make their own decisions based on a full understanding of the risks associated with infectious diseases and the advantages/disadvantages of being vaccinated. They need to be able to obtain health information to inform their decisions regarding vaccination against emerging infectious diseases that may cause global pandemics. This is important because the COVID-19 vaccine is the first mRNA vaccine to be used worldwide.

There are various sources of information about vaccines, including TV, radio, newspapers, the Internet (e.g., websites, social networking services), friends/colleagues, and medical institutions/medical personnel. Workers who responded that they “trusted” these sources of information were significantly more likely to want to be vaccinated against COVID-19 [8]. Conversely, it has been reported that those who obtained information about the vaccine from social networking service were less likely to want to be vaccinated [9]. Although the Internet offers the advantage of sharing a large amount of information quickly, it also has potential to spread false information, disinformation, and rumors during health emergencies, which hinders effective responses and creates confusion and mistrust [10]. The World Health Organization used the term “infodemic” to describe this situation. Infodemics, mainly disseminated through the Internet, including social networking services, have been reported to increase vaccine hesitancy [11-13]. However, no studies have clarified the impact of different sources of health information on people who are not willing to receive the COVID-19 vaccine.

The purpose of this study was to clarify the relationship between sources of health information and willingness to receive the COVID-19 vaccine among those who were not willing to get the vaccine as at the end of December 2020, which was during the third wave of infection in Japan.

## Methods

### Study design and participants

This prospective cohort study was conducted by a research group from the University of Occupational and Environmental Health, Japan. The project was named the Collaborative Online Research on Novel-coronavirus and Work (CORoNaWork) study. Data were collected using a self-administered online questionnaire survey delivered via the Internet survey company Cross Marketing Inc. (Tokyo, Japan). The baseline survey was conducted on December 22–25, 2020, and the follow-up survey on February 18–19, 2021. In both periods, Japan was in the third wave of the COVID-19 pandemic, where the number of COVID-19 infections and deaths was markedly higher than in the first and second waves.

Details of the study protocol, including the sampling plan and participant recruitment procedure, have been previously reported [14]. Participants were aged 20–65 years and worked at the time of the baseline survey (N = 33,087). CORoNaWork study participants were stratified by sex, age, and region of residence using cluster sampling. After excluding 6,051 initial participants who provided invalid responses, we included 27,036 participants in the study database. These participants received a follow-up survey, to which 19,941 people responded (74% follow-up rate). We excluded 7,415 participants who answered “yes” to the question “If a COVID-19 vaccine becomes available, would you like to get it?” in the baseline survey. We finally analyzed responses for 12,526 participants. Figure1 shows the flow diagram for this study.

The present study was approved by the Ethics Committee of the University of Occupational and Environmental Health, Japan (Approval number: R2-079 and R3-006). Informed consent was obtained from all participants.

**Figre1.** Flow diagram of this study

### Assessment of intention to receive the COVID-19 vaccination

In the follow-up survey, we asked participants, “Do you want to get the COVID-19 vaccine?” Participants could choose one of four response options: “I want to,” “I want to a little,” “I do not really want to,” and “I do not want to.” Those who chose “I want to” or “I want to a little” were classified as being willing to receive the COVID-19 vaccine; other participants were classified as not willing.

### Source of health information

In the follow-up survey, we asked participants, “Which medium do you use most to get information to protect your health?” Participants could choose one of seven options: TV; Internet; newspaper; radio; magazines; friends/colleges; and others.

### Assessment of covariates

Covariates included demographics, socioeconomic factors, occupation, industry, and fear of COVID-19 transmission. Age was classified into five age groups: 20–29, 30–39, 40–49, 50–59, and 60–65 years. Education was classified into three categories: junior high or high school, vocational school or college, and university or graduate school. Yearly household income was classified into four categories: <2.50 million Japanese yen (JPY), 2.50–3.74 million JPY, 3.75–5.24 million JPY, and ≥5.25 million JPY. We classified occupation into 10 categories: general employee; manager; executive manager; public employee, faculty member, or non-profit organization employee; temporary or contract employee; self-employed; small office/home office; agriculture, forestry, or fishing; professional occupation (e.g., lawyer, tax accountant, medical-related); and other occupations. Participants could choose one of 22 options for their work industry: energy, materials, industrial machinery; food; beverages/tobacco products; pharmaceuticals/medical supplies; cosmetics/toiletries/sanitary products; fashion and accessories; precision machinery and office supplies; home appliances/audio visual equipment; automobiles and transportation equipment; household goods; hobby/sporting goods; real estate and housing equipment; information and communication; wholesale and retail; finance/insurance; transportation and leisure; restaurant and other services; public offices and organizations; education, medical services, religion; mass media; market research; and others. These industries were then classified into nine categories based on the International Standard Industrial Classification of All Economic Activities: manufacturing; public service; information and communication; wholesale and retail; food service; medical and welfare; finance and insurance; construction; and others. To evaluate fear of COVID-19 transmission, we asked participants, “Do you feel anxious about getting infected with COVID-19?” (Yes or No).

The cumulative incidence rate of COVID-19 infection in the prefecture of residence one month before the baseline survey was used as a community-level variable.

### Statistical analyses

Multilevel logistic regression analyses were used to examine the associations between the source of health information and willingness to receive the COVID-19 vaccine. Age-sex adjusted odds ratios (ORs) and multivariate adjusted ORs were estimated with a multilevel logistic model nested in the prefecture of residence. The multivariate model was adjusted for age, sex, educational background, income, occupation, industry, and fear of COVID-19 transmission. A p-value less than 0.05 was considered statistically significant. All analyses were conducted using Stata (Stata Statistical Software release 16; StataCorp LLC, TX, USA).

## Results

Table 1 shows participants’ characteristics by the source of health information. The Internet was the most popular source of information, followed by TV, newspapers, and friends/colleagues. There was almost no difference between men and women for TV and the Internet, but newspapers, radio, and magazines were mostly used by men, whereas friends/colleagues were mostly used by women. Those who used the Internet, magazines, and friends/colleagues as information sources were more likely to be younger, whereas those who used TV, newspapers, and radio were more likely to be older. Those who used TV for information were more likely to be willing to receive the COVID-19 vaccine than those who used the Internet, newspapers, and friends/colleagues.

**Table 1.** Characteristics of the study population by information source

|  | TV | Internet | Newspaper | Radio | Friends and colleagues | Other |
| --- | --- | --- | --- | --- | --- | --- |
|  | n(%) | n(%) | n(%) | n(%) | n(%) | n(%) |
| Total | 3586 | 7299 | 522 | 137 | 396 | 586 |
| Sex |  |  |  |  |  |  |
| Men | 1810 (50.5) | 3756 (51.5) | 366 (70.1) | 101 (73.7) | 152 (38.4) | 348 (59.4) |
| Age (years) |  |  |  |  |  |  |
| 20–29 | 149 (4.2) | 477 (6.5) | 9 (1.7) | 1 (0.7) | 30 (7.6) | 36 (6.1) |
| 30–39 | 523 (14.6) | 1419 (19.4) | 43 (8.2) | 14 (10.2) | 72 (18.2) | 111 (18.9) |
| 40–49 | 1029 (28.7) | 2404 (32.9) | 150 (28.7) | 39 (28.5) | 135 (34.1) | 170 (29.0) |
| 50–59 | 1386 (38.7) | 2301 (31.5) | 222 (42.5) | 58 (42.3) | 117 (29.5) | 215 (36.7) |
| 60–65 | 499 (13.9) | 698 (9.6) | 98 (18.8) | 25 (18.2) | 42 (10.6) | 54 (9.2) |
| Education |  |  |  |  |  |  |
| Junior high or high school | 1081 (30.1) | 1850 (25.3) | 121 (23.2) | 38 (27.7) | 108 (27.3) | 191 (32.6) |
| Vocational school or college | 930 (25.9) | 1673 (22.9) | 87 (16.7) | 28 (20.4) | 116 (29.3) | 124 (21.2) |
| University or graduate school | 1575 (43.9) | 3776 (51.7) | 314 (60.2) | 71 (51.8) | 172 (43.4) | 271 (46.2) |
| Annually household income (million JPY)□ |  |  |  |  |  |  |
| <2.00 | 270 (7.5) | 500 (6.9) | 29 (5.6) | 21 (15.3) | 28 (7.1) | 70 (11.9) |
| 2.00–3.99 | 800 (22.3) | 1493 (20.5) | 79 (15.1) | 29 (21.2) | 95 (24.0) | 132 (22.5) |
| 4.00–5.99 | 866 (24.1) | 1804 (24.7) | 117 (22.4) | 33 (24.1) | 86 (21.7) | 131 (22.4) |
| 6.00–7.99 | 673 (18.8) | 1458 (20.0) | 120 (23.0) | 20 (14.6) | 79 (19.9) | 113 (19.3) |
| 8.00–9.99 | 470 (13.1) | 894 (12.2) | 69 (13.2) | 17 (12.4) | 41 (10.4) | 56 (9.6) |
| ≥ 10.00 | 507 (14.1) | 1150 (15.8) | 108 (20.7%) | 17 (12.4) | 67 (16.9) | 84 (14.3) |
| Occupation |  |  |  |  |  |  |
| General employee | 1620 (45.2) | 3452 (47.3) | 180 (34.5) | 70 (51.1) | 196 (49.5) | 250 (42.7) |
| Manager | 311 (8.7) | 675 (9.2) | 69 (13.2) | 10 (7.3) | 20 (5.1) | 45 (7.7) |
| Executive manager | 100 (2.8) | 216 (3.0) | 29 (5.6) | 3 (2.2) | 9 (2.3) | 17 (2.9) |
| Public employee, faculty member, or non-profit organization employee | 341 (9.5) | 711 (9.7) | 85 (16.3) | 15 (10.9) | 35 (8.8) | 42 (7.2) |
| Temporary contract employee | 439 (12.2) | 763 (10.5) | 50 (9.6) | 12 (8.8) | 35 (8.8) | 59 (10.1) |
| Self employed | 382 (10.7) | 687 (9.4) | 53 (10.2) | 17 (12.4) | 43 (10.9) | 69 (11.8) |
| SOHO | 59 (1.6) | 141 (1.9) | 5 (1.0) | 0 (0.0) | 4 (1.0) | 12 (2.0) |
| Agriculture, forestry and fishery | 30 (0.8) | 63 (0.9) | 3 (0.6) | 0 (0.0) | 3 (0.8) | 11 (1.9) |
| Professional occupation (e.g., lawyer, tax accountant, medical-related) | 211 (5.9) | 393 (5.4) | 34 (6.5) | 3 (2.2) | 34 (8.6) | 48 (8.2) |
| Other occupation | 93 (2.6) | 198 (2.7) | 14 (2.7) | 7 (5.1) | 17 (4.3) | 33 (5.6) |
| Industry category |  |  |  |  |  |  |
| Manufacturing | 591 (16.5) | 1246 (17.1) | 82 (15.7) | 36 (26.3) | 62 (15.7) | 78 (13.3) |
| Public service | 243 (6.8) | 456 (6.2) | 54 (10.3) | 7 (5.1) | 23 (5.8) | 35 (6.0) |
| Information and technology | 185 (5.2) | 443 (6.1) | 22 (4.2) | 6 (4.4) | 10 (2.5) | 32 (5.5) |
| Retail and wholesale | 267 (7.4) | 501 (6.9) | 35 (6.7) | 7 (5.1) | 24 (6.1) | 34 (5.8) |
| Eating/drinking | 204 (5.7) | 381 (5.2) | 16 (3.1) | 7 (5.1) | 31 (7.8) | 24 (4.1) |
| Medical and welfare | 521 (14.5) | 1077 (14.8) | 89 (17.0) | 17 (12.4) | 90 (22.7) | 94 (16.4) |
| Finance | 177 (4.9) | 300 (4.1) | 25 (4.8) | 5 (3.6) | 15 (3.8) | 22 (3.8) |
| Construction | 144 (4.0) | 276 (3.8) | 25 (4.8) | 5 (3.6) | 13 (3.3) | 11 (1.9) |
| Other | 1254 (35.0) | 2619 (35.9) | 174 (33.3) | 47 (34.3) | 128 (32.3) | 254 (43.3) |
| Fear of COVID-19 transmission |  |  |  |  |  |  |
| Yes | 2974 (82.9) | 5702 (78.1) | 393 (75.3) | 96 (70.1) | 291 (73.5) | 328 (56.0) |
| Willingness to get the COVID-19 vaccine |  |  |  |  |  |  |
| Yes | 2033 (56.7) | 3646 (50.0) | 278 (53.3) | 67 (48.9) | 168 (42.4) | 201 (34.3) |

JPY, Japanese yen; SOHO, small office/home office.

Table 2 shows the associations between the source of health information and willingness to receive the COVID-19 vaccine. The age and sex adjusted analysis showed that compared with TV, significantly lower willingness to be vaccinated was associated with using the Internet (OR=0.78, 95% confidence interval [CI]: 0.72–0.85, p<0.001), newspapers (OR=0.82, 95% CI: 0.68–0.99, p=0.037), radio (OR=0.69, 95% CI: 0.49–0.97, p=0.033), and friends/colleagues (OR=0.58, 95% CI: 0.47–0.72, p<0.001) as sources of information. After adjusting for demographics, including socioeconomic factors, occupation, industry, and fear of COVID-19 transmission, associations between willingness to be vaccinated and using the Internet (OR=0.81, 95% CI: 0.74–0.88, p<0.001) and friends/colleagues (OR=0.62, 95% CI: 0.50–0.77, p<0.001) as information sources were still significantly lower than TV.

**Table 2.** Association between source of health information and willingness to receive the COVID-19 vaccine

| Source of health information | Sex and age adjusted |  |  |  | Multivariate adjusted* |  |  |  |
| --- | --- | --- | --- | --- | --- | --- | --- | --- |
|  | OR | 95% CI |  | P value | OR | 95% CI |  | P-value |
| TV | reference |  |  |  | reference |  |  |  |
| Internet | 0.78 | 0.72 | 0.85 | <0.001 | 0.81 | 0.74 | 0.88 | <0.001 |
| Newspaper | 0.82 | 0.68 | 0.99 | 0.037 | 0.83 | 0.68 | 1.00 | 0.056 |
| Radio | 0.69 | 0.49 | 0.97 | 0.033 | 0.78 | 0.55 | 1.12 | 0.181 |
| Friends and colleagues | 0.58 | 0.47 | 0.72 | <0.001 | 0.62 | 0.50 | 0.77 | <0.001 |
| Other | 0.40 | 0.33 | 0.48 | <0.001 | 0.51 | 0.42 | 0.62 | <0.001 |

\* Adjusted for sex, age, education, income, occupation, industry, and fear of COVID-19 transmission.
OR, odds ratio; CI, confidence interval.

## Discussion

The OR for the change in willingness to be vaccinated from “no” to “yes” differed by source of health information. Compared with workers who reported using TV as a source of information, those who reported getting information from the Internet and friends/colleagues had significantly lower ORs for willingness to receive the COVID-19 vaccine.

Google publishes a relative percentage of the number of searches per week going back 1 year from the search date, with the highest value for that period being 100. At the time of the search for this study (June 29, 2021), the highest number of searches for “vaccine” was during June 13–19, 2021. The number of searches during that period was 100, whereas the number of searches at the time of the first survey was 9, but increased to 28 at the time of the second survey [15]. This indicated that many Internet users were actively trying to find information about vaccines during this time. When searching for online health information on the Internet, people tend to select information that confirms their existing beliefs; this introduces the risk of confirmation bias, which results in a biased evaluation of the information [16]. In addition, because the information displayed changes depending on the search history, the information obtained is likely to be biased. If one tries to obtain negative information about vaccines from the outset, it is likely that one may acquire false information arising from the infodemic, and then continue to acquire similar information. This may result in increasing vaccine hesitancy.

Social networking services are an Internet-based source of information, but they have different characteristics from websites. When people obtain information from others via a social networking service, they tend to form groups with similar systems of beliefs. This is because of the echo chamber effect, whereby an individual’s opinions are amplified and strengthened by continuously seeing and hearing similar opinions [17, 18]. Therefore, if people mainly obtain health information from social networking services, they may be strengthening their original vaccine hesitancy.

It has been reported that whether friends are vaccinated influences an individual’s willingness to be vaccinated; if none of an individual’s friends are vaccinated, it has a negative effect [19]. As the second survey for this study was conducted just after the vaccination of healthcare workers started, it is assumed that few friends/colleagues of our participants had been vaccinated. Their willingness to get the vaccine may therefore not have increased if they mainly received information from their friends or colleagues. Vaccination started in many areas after the second survey for this study, and people’s willingness to get the vaccine may therefore have changed according to the vaccination status of surrounding friends.

The results of this study were consistent with those of other studies that found willingness to be vaccinated was higher in those that used TV as a source of health information^11)^. Information can be obtained passively from TV, and is widely disseminated to the indifferent population regardless of whether they have vaccine hesitancy. During the period of the second survey, there was an increase in TV coverage regarding vaccines because vaccination of healthcare workers had just started. Therefore, it is possible that the group that mainly received health information from TV changed from not wanting to be vaccinated to wanting vaccination.

Willingness to receive the vaccine has been identified as a crucial issue in the development of vaccination programs [20]. It is important that the approach to vaccine hesitancy is not uniform, and it is necessary to listen to the concerns of those with low vaccination willingness in their individual contexts and implement appropriate communication [21]. As this survey targeted workers, it is also important to implement workplace-based approaches to increase willingness to be vaccinated and improve the vaccination rate. When implementing vaccination programs in the workplace, it is necessary to understand that different sources of health information may result in different attitudes toward vaccination, and to promote awareness of vaccination programs. Individual concerns can be clarified by confirming usual information sources, content, and vaccination preferences with the target population. Changes in vaccination willingness can be expected when occupational health professionals interview those who do not want to be vaccinated and provide guidance focused on their concerns. Routine provision of health-related information by occupational health professionals may enhance workers’ health literacy and information choices, decisions, and actions. Furthermore, actively disseminating correct information via TV, websites, social networking services, other media (e.g., YouTube), and by occupational health professionals in the workplace may expand the options for correct information.

Regarding organizational factors and vaccination behavior in the workplace, it has been reported that organizational policies on vaccination can influence vaccination rates [22]. Therefore, when implementing a vaccination program for workers, a message from the top management of that company about vaccination and creating an environment that facilitates access to vaccination is recommended to protect workers’ health and may increase the vaccination rate. It is also known that in workplaces where perceived organizational support exists, there is a sense of responsibility to the organization and altruistic behavior [23]. This means that employees will act to achieve group immunity to protect colleagues, which suggests that willingness to be vaccinated will increase. Therefore, daily health promotion activities that increase perceived organizational support may also contribute to improving vaccination rates.

There were several limitations of this study that should be noted. First, the survey was conducted via an Internet panel. Because of the bias of the target population (a population with Internet access), the generalizability of the results is limited. Second, the timing of the survey might have impacted our results. In Japan, vaccination against COVID-19 began on February 17, 2021, for healthcare workers, in April 2021 for people aged ≥65 years, and on June 21, 2021, for people in workplaces. Because the trends in vaccination changed rapidly, careful consideration is needed to determine whether the present findings apply to all periods. Third, the causal relationship between the source of information and willingness to get the vaccine was not clear. But because this was a prospective cohort study, the influence of the temporal relationship between predictors and outcomes is likely to be strong. Finally, there is a common method bias. Further studies using objective records of whether people were actually vaccinated are needed.

## Conclusions

This study suggests there is a difference in the change in willingness to receive the COVID-19 vaccine depending on the source of health information. Compared with the group that mainly obtains information from TV, those that obtain information from the Internet and friends/colleagues did not show increased willingness to be vaccinated. It is important to reach out to workers who do not watch TV when implementing vaccination programs in the workplace. It can be expected that the willingness to vaccinate people who do not wish to be vaccinated can be changed by an active policy whereby the top management of a company recommend vaccination and implement an individual approach by occupational health professionals.

## Data Availability

For availability of data and material please contact the corresponding author.

## Abbreviations

COVID-19: coronavirus disease 2019
JPY: Japanese yen
OR: odds ratio
CI: confidence interval

## Declarations

### Ethical approval and consent to participate

This study was approved by the Ethics Committee of the University of Occupational and Environmental Health, Japan (reference No. R2-079 and R3-006). Informed consent was obtained from all participants via the survey website.

### Consent for publication

Not applicable.

### Availability of data and material

For availability of data and material please contact the corresponding author.

## Authors’ contributions

KH; analysis and writing the manuscript, TN and TM; creating the questionnaire, analysis, drafting the manuscript and advice on interpretation, HA, AH, ST, MT, and SM; Review of manuscripts, and advice on interpretation, YF; overall survey planning, creating the questionnaire, and review of manuscripts, and advice on interpretation.

## Acknowledgements

The current members of the CORoNaWork Project, in alphabetical order, are as follows: Dr. Yoshihisa Fujino (present chairperson of the study group), Dr. Akira Ogami, Dr. Arisa Harada, Dr. Ayako Hino, Dr. Hajime Ando, Dr. Hisashi Eguchi, Dr. Kazunori Ikegami, Dr. Kei Tokutsu, Dr. Keiji Muramatsu, Dr. Koji Mori, Dr. Kosuke Mafune, Dr. Kyoko Kitagawa, Dr. Masako Nagata, Dr. Mayumi Tsuji, Ms. Ning Liu, Dr. Rie Tanaka, Dr. Ryutaro Matsugaki, Dr. Seiichiro Tateishi, Dr. Shinya Matsuda, Dr. Tomohiro Ishimaru, and Dr. Tomohisa Nagata. All members are affiliated with the University of Occupational and Environmental Health, Japan.

